# Impact of COVID-19 on Pediatric Emergency Department Visits: A Retrospective Cohort Study

**DOI:** 10.1101/2021.04.05.21254921

**Authors:** Pierre Fontaine, Esli Osmanlliu, Jocelyn Gravel, Ariane Boutin, Evelyne D. Trottier, Nathalie Orr Gaucher, Antonio D’Angelo, Olivier Drouin

## Abstract

**Background & Objective:** COVID-19 has caused significant shifts in healthcare utilization, including pediatric emergency departments (EDs). We describe variations in visits made to two large pediatric EDs during the first three months of the COVID-19 pandemic, compared to a historical control period.

**Methods:** We performed a retrospective cohort study of children presenting to two academic pediatric EDs in Quebec, Canada. We compared the number of ED visits during the first wave of COVID-19 pandemic (March-May 2020) to historical controls (March-May 2015-2019), using Poisson regression, adjusting for site and the underlying baseline trend. Secondary analyses examined variations in ED visits by acuity, disposition, and disease categories.

**Results:** From 2015 to 2019, the two EDs had a median of 1,632 visits per week [interquartile range (IQR) 1,548; 1,703]; in 2020, this number decreased to 536 visits per week [IQR 446; 744]. In multivariable analyses, this represent a 53.3% (95%CI: 52.1, 54.4) reduction in the number of ED visits. The reduction was larger among visits triage categories 4 and 5 (lower acuity) than categories 1, 2 and 3 (higher acuity): -54.2% vs. -42.0% (p<0.001). A greater proportion of children presenting to these sites were hospitalized during the COVID period than in pre-COVID period: 11.8% vs. 5.5% (p<0.001).

**Conclusions:** During the early stages of the COVID-19 pandemic, there was a large decrease in visits to pediatric EDs. Patients presented with higher acuity at triage and the proportion of patients requiring hospitalization increased.

## Introduction

As of December 2020, the novel coronavirus (SARS-CoV-2) has caused over 73 million infections and 1,6 million deaths worldwide. [1] During the same period, the province of Québec, Canada, registered over 167,000 cases and 7,570 deaths. [2] Beyond the direct effects of the virus, public health measures enacted to contain the virus have had broad ranging collateral effects. In parallel, there have been significant declines in adult Emergency Department (ED) visits during the early stages of the pandemic. [3] This included decreases in ED visits for acute illnesses unrelated to COVID-19 or other infectious causes that would normally require rapid medical attention, [4] such as strokes, urolithiasis, and abdominal pain. [5, 6]

In the pediatric population, clinical manifestations of COVID-19 are generally less severe than in adults, and typically have a more favorable clinical course. [7-11] Like in adults, there have been reports of decreases in pediatric ED care use internationally. [12, 13] For example, an early report from Italy showed a significant reduction in pediatric ED visits [14] and researchers in Germany reported a 64% decrease in pediatric ED utilization during the COVID-19 lockdown. [15] A study from Philadelphia demonstrated a 67% decrease in the mean number of daily pediatric ED visits, while the proportion of patients triaged as high acuity increased after the state-wide stay-at-home order. [16]

Nonetheless, a knowledge gap remains regarding the impact of the pandemic on pediatric ED visits in Canada and evidence on resource utilization in order to inform care delivery re-organization as the pandemic continues. The main objective of this study was to document changes in the number of visits during the first wave of the COVID-19 pandemic by comparing the number of ED visits from March to May 2020 to a historical control period (2015-2019) in two Canadian pediatric EDs. Our secondary objective was to determine if the acuity and disease categories of ED visits differed between the two study periods.

## Methods

### Study Design and Setting

We conducted a multicentric retrospective cohort study using the ED information management systems of the CHU Sainte-Justine (CHUSJ) and Montreal Children’s Hospital (MCH), two large tertiary care, university-affiliated, pediatric hospitals in Montreal, Canada, each with an annual census of more than 75 000 patients-visits per year each. Both hospitals receive COVID-19 cases, and since the beginning of the pandemic, the CHUSJ has received transfer from across the province for positive pediatric cases requiring hospitalization.

### Participants

Patients included in the study were all those younger than 18 years of age who were evaluated by a physician in either of the two EDs during the study periods (i.e., between March 1st and May 31st in the years 2015, 2016, 2017, 2018, 2019, and 2020). Patients who left the ED before being seen by a physician were excluded from the primary analysis but included in secondary analyses.

### Outcomes

The primary outcome was the overall number of weekly ED visits during the study period. Secondary outcomes included the percent decrease in the number of visits between study periods.

### Exposure

The main exposure of interest was the period during which the patient was seen: pre-COVID period vs. COVID period. In bivariate analyses, study periods were divided in weeks to allow comparison between years: pre-COVID period included weeks 10-20 (March-May) of 2015-2019, whereas the COVID period covered weeks 10-20 (March-May) of 2020. This accounted for a total of 55 weeks per hospital during the pre-COVID period, and 11 weeks per hospital during the COVID period.

### Covariates

Covariates included in the analysis were hospital (CHUSJ vs. MCH), age category of the patient (<1 year old, 1-4, 5-11, and 12-17 years of age), triage level according to the Canadian Triage and Acuity Scale (CTAS; ranging from level 1 [patient requiring resuscitation] to level 5 [non-urgent visit]), [17] time (shift) of the visit (day, evening, or night), and ED disposition plan. ICD-10 diagnostic categories [18] at discharge were only available from one of the study sites (MCH). The distribution of select frequent ICD-10 diagnostic categories representing common reasons for pediatric EDs visits is presented (Appendix A).

### Analysis

The primary analysis examined the association between study period (pre-COVID vs. COVID period) and the weekly number of ED visits. This was conducted first in a bivariate analysis using the Mann-Whitney U test, then in a multivariate Poisson regression model, adjusting for hospital and baseline trend. The same analyses were subsequently used to estimate the impact of the COVID period on other covariates: patient age categories, CTAS levels, shift of the visit, and ED disposition during the pre-COVID vs. COVID period. Following a Bonferroni adjustment, the level of statistical significance (alpha) was set to a p-value < 0.00172 (0.05/29 comparisons). All analyses were conducted using R (Version 4.0.3, The R Foundation for Statistical Computing) and R Studio (Version 1.2.1335, RStudio, Inc.). This study was approved by the ethics review board of CHUSJ on July 28^th^, 2020. Consent was waived by the institution’s review board given the use of an administrative de-identified database.

## Results

There were a total of 178,830 visits made during the pre-COVID period, and 14,628 visits during the COVID period. Age distribution and visit characteristics during both study periods are shown in Table 1. Across both hospitals, the median number of weekly visits decreased from 1632 in the pre-COVID period to 536 in the COVID period (p < 0.001) (Table 1, Figure 1a). Week 14 (March 30^th^ to April 4^th^) of 2020, marked the lowest number of weekly visits at each site (439 for CHUSJ; 335 for MCH). Using a multivariable Poisson regression model allowing adjustment for hospital and baseline trend, the reduction in weekly visits during the COVID period was 889 (95%CI: 776, 1002) visits, which corresponds to a decrease of 53.3% (95%CI: 52.1, 54.4).

**Table 1.**
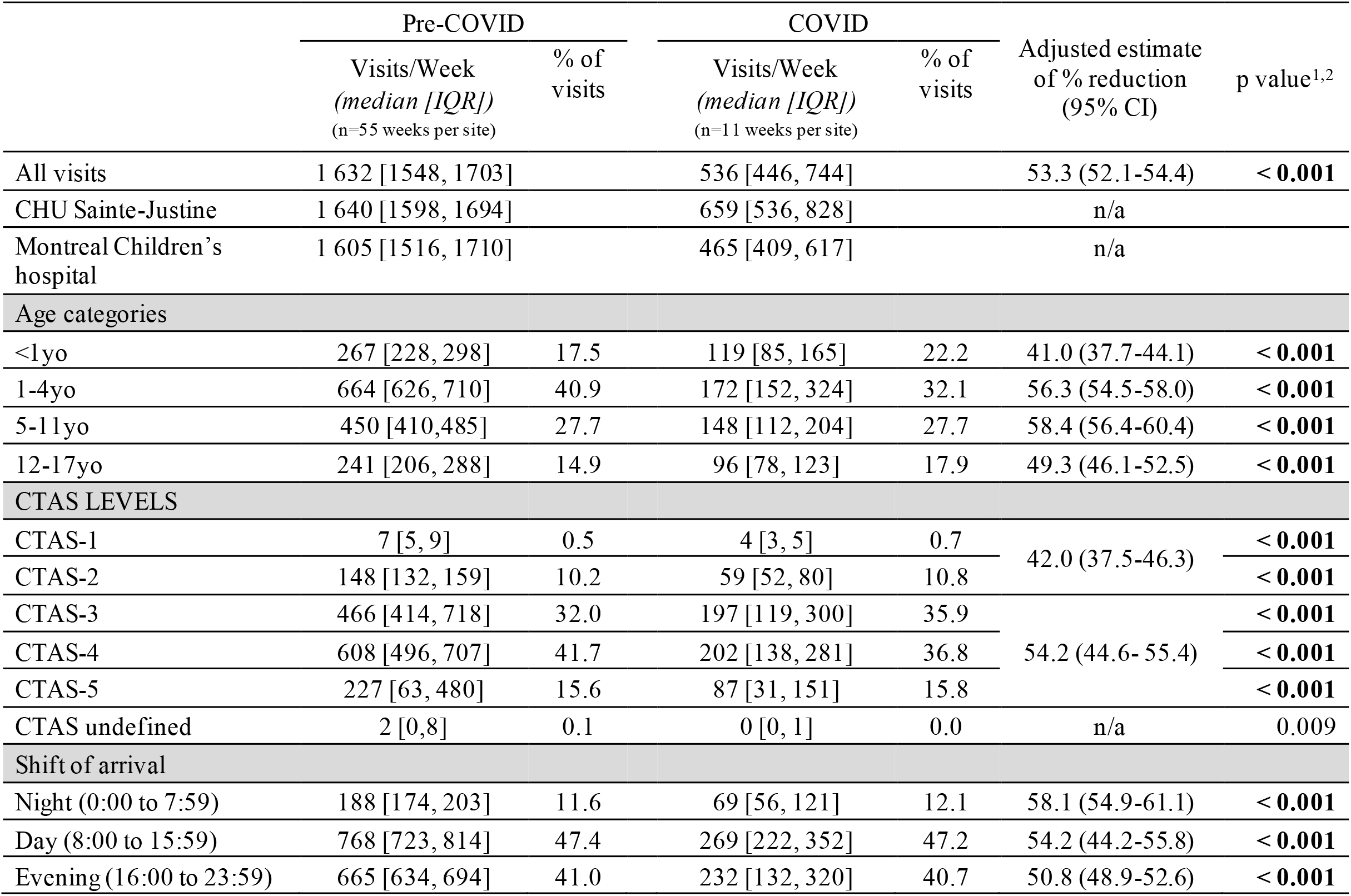
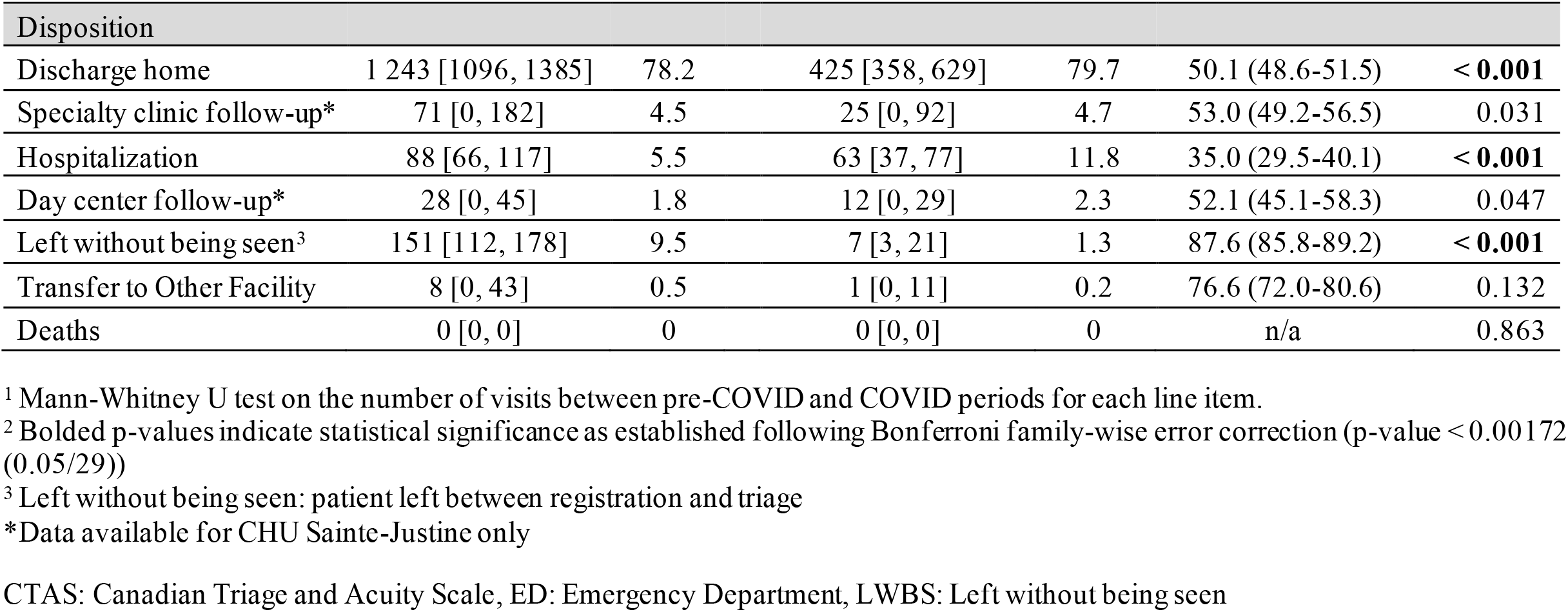
Median (Interquartile range) weekly visits to two Quebec Pediatric EDs, pre-COVID vs. COVID periods

**Figure 1:**
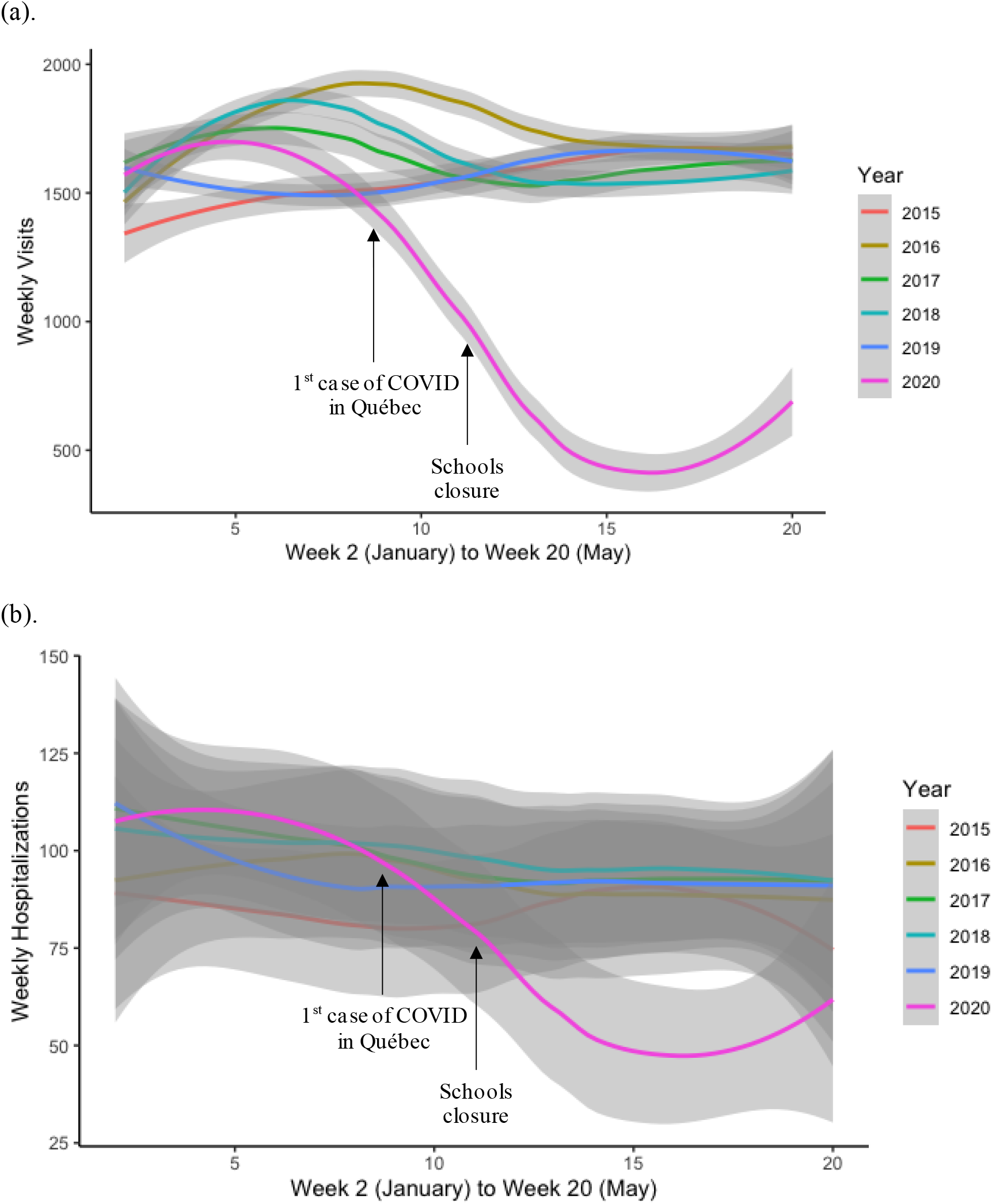
(a)Total Weekly Visits From 2015 to 2020 at the two Pediatric EDs in Quebec and (b) Weekly Number of Hospitalizations From 2015 to 2020 at the two Pediatric EDs in Quebec *Shaded area corresponds to the 95% confidence interval*.

In bivariate analyses, there was a significant difference in the distribution of triage levels (CTAS) between the pre-COVID vs. COVID period (p < 0.001) (Table 1). Stratifying by triage levels, for CTAS 1-2, there was a decrease from a median of 156 [IQR 140; 168] visits/week in pre-COVID compared to 62 [IQR 54; 83] visits/week during the COVID period. This represents a 42.0% (95%CI: 37.5, 46.3) decrease in the number of visits (Figure 2a. p < 0.001). For CTAS 3-5, the median number of weekly visits decreased from 1463 [IQR 1395; 1518] pre-COVID to 474 [IQR 390; 668] in the COVID period, a 54.2% (95%CI 52.4, 55.4) decrease (Figure 2b. p<0.001).

**Figure 2:**
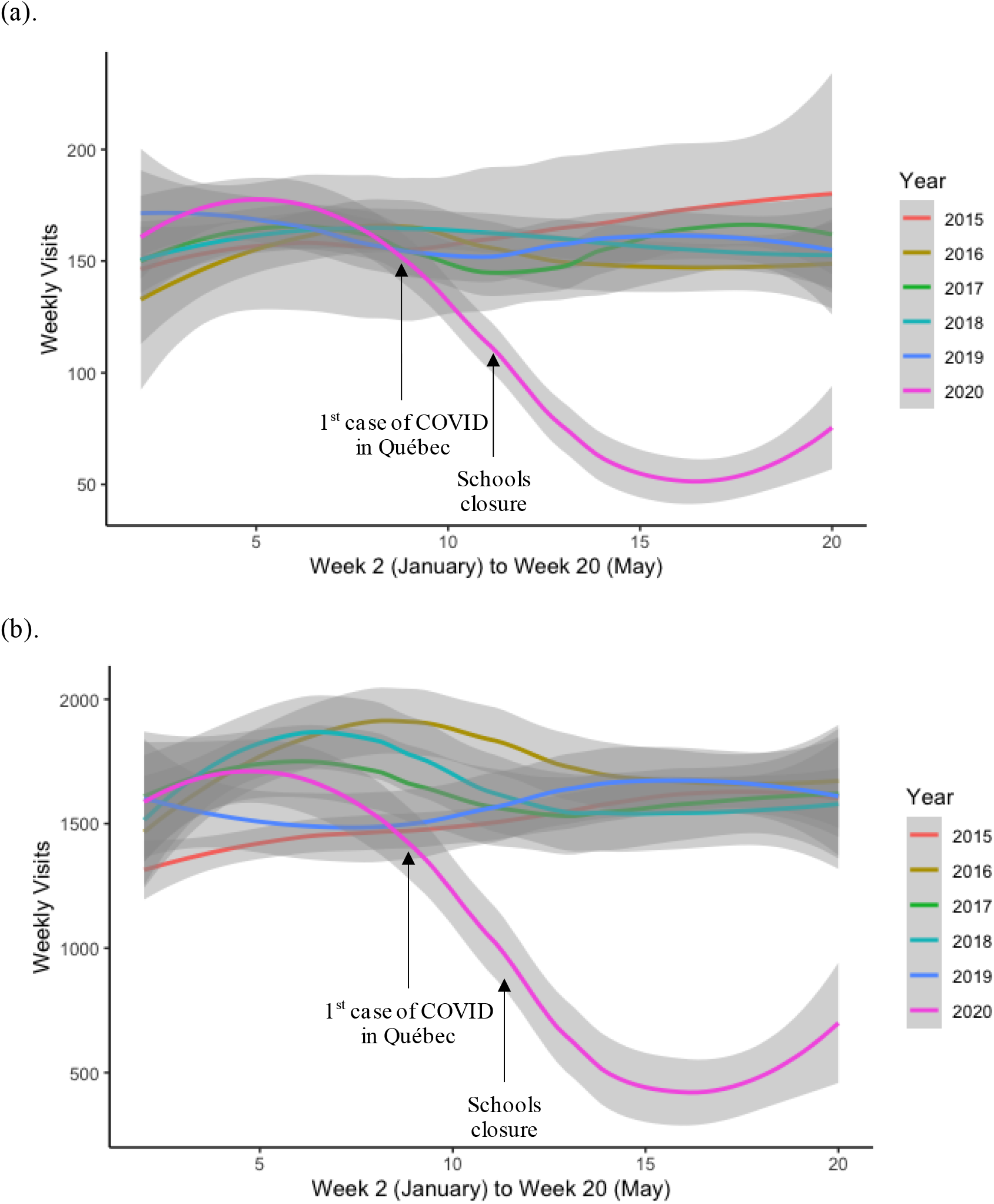
(a) CTAS 1-2 Weekly Visits From 2015 to 2020 at the two Pediatric EDs in Quebec and (b) CTAS 3-5 Weekly Visits From 2015 to 2020 at the two Pediatric EDs in Quebec *Shaded area corresponds to the 95% confidence interval*.

The median number of ED visits resulting in hospitalization decreased from 88 per week pre-COVID to 63 per week during COVID, a 35.0% (95%CI 29.5, 40.1) decrease (Figure 1b). However, the proportion of ED visits resulting in hospitalization increased during the COVID period relative to pre-COVID: 11.8% vs. 5.5%, (p<0.001). With regards to disease categories, the included diagnostic categories capture 65.5% and 53.3% of all visits in the pre-COVID and COVID periods, respectively (Table 2). The absolute number of ED visits in each category decreased during the COVID period compared to pre-COVID period (Table 2). When examined as a proportion of visits however, the only disease categories that showed statistically significant changes were *Fever unspecified, viral infection, URTI, Bronchiolitis* (25.1% pre-COVID vs 20.5% COVID period, p<0.001), *Diseases of the respiratory system* (19.0% pre-COVID vs. 10.5% COVID period, p<0.001), and *Diseases of the eye, adnexa, ear, and mastoid process* (7.2% vs. 3.2%, p<0.001) (Table 2). Conversely, there was a significant increase in the proportion of injuries (8.2% pre-COVID vs. 12.4% COVID period, p< 0.001), and *Diseases of the digestive system* (2.8% vs. 4.3%, p= 0.002). Finally, in a secondary analysis, the number of patients who left without being seen (LWBS) decreased during the COVID period compared to pre-COVID (9.5% vs. 1.3%., p<0.001).

**Table 2.** Distribution of the most common ICD-10 diagnostic categories at MCH ED discharge, pre-COVID vs. COVID^1^

|  | Pre-COVID |  | COVID |  | Adjusted estimate of % reduction (95% IC) | p value<sup>2,3</sup> |
| --- | --- | --- | --- | --- | --- | --- |
|  | Visits/Week<br>(median [IQR])<br>(n=55 weeks) | Proportion of<br>MCH weekly<br>visits (%) | Visits/Week<br>(median [IQR])<br>(n=11 weeks) | Proportion of<br>MCH weekly<br>visits (%) |  |  |
| *Fever unspecified, viral infection, URTI, Bronchiolitis* | 373 [330, 400] | 25.1 | 96 [78, 215] | 20.5 | 51.1 (48.2-53.8) | **< 0.001** |
| *Diseases of the respiratory system* | 279 [257, 311] | 19.0 | 49 [34, 164] | 10.5 | 60.4 (57.5-63.0) | **< 0.001** |
| *Injuries* | 134 [124, 163] | 8.2 | 58 [45, 78] | 12.4 | 54.5 (50.2-58.5) | **< 0.001** |
| *Diseases of the eye and adnexa, ear, and mastoid process* | 109 [104, 130] | 7.2 | 15 [14, 44] | 3.2 | 65.4 (60.9-69.4) | **< 0.001** |
| *Diseases of the digestive system* | 47 [41, 51] | 2.8 | 20 [19, 27] | 4.3 | 50.3 (42.0-57.4) | **< 0.002** |
| *Diseases of the nervous system* | 17 [12, 20] | 1.0 | 5 [3, 7] | 1.1 | 61.6 (49.5-71.1) | 0.016 |
| *Mental health or substance abuse related* | 13 [11, 16] | 0.8 | 4 [2, 6] | 0.9 | 66.4 (55.0-75.2) | 0.029 |
| *Endocrine, nutritional, and metabolic diseases* | 5 [4, 6] | 0.2 | 1 [1, 3] | 0.2 | 64.1 (41.9-78.6) | n/a |
| *Diseases of the circulatory system* | 4 [2, 5] | 0.2 | 1 [1, 3] | 0.2 | 23.5 (0.0-56.1) | n/a |
| *Other diagnoses* | n/a | 35.5 | n/a | 46.7 | n/a | n/a |

<sup>1</sup> ICD-10 diagnostic categories at discharge were only available from one of the study sites (MCH).
<sup>2</sup> Mann-Whitney U test on the number of visits between pre-COVID and COVID periods for each line item.
<sup>3</sup> Bolded p-values indicate statistical significance as established following Bonferroni family-wise error correction (p-value < 0. 172 (0.05/29))

## Discussion

This retrospective study shows a 53.3% decrease in visits made to two pediatric EDs visits during the early stage of the COVID-19 pandemic (March-May 2020) compared to historical control. These findings are consistent with other reports showing reductions in ED visits of 50%-88% compared to pre-COVID. [12-16, 19]

Our results also show a reduction in the number, but increase in proportion of high acuity patients as measured by the CTAS during the early COVID period relative to the pre-COVID period. Indeed, while there was a decrease in average weekly visits for each triage category, this decrease was greater for visits with triage levels 3, 4 and 5 than levels 1 and 2 (decrease of 54.2% and 42.0% respectively). We also saw an increase in the proportion of visits requiring hospitalization. This shift to higher acuity visits during COVID-19 is similar to studies conducted in other pediatric EDs. [16]

One possible explanation for the large observed decrease in the number of visits, and the relative increase in the acuity of visits, is that public health measures issued by authorities to reduce the spread of COVID-19 likely reduced the rate of transmission of other infectious diseases such as upper respiratory tract infections and diseases of the respiratory system which usually represent the bulk of the visits during this period of the year. Given the observed decreases in visit numbers even for high acuity triage levels, it is also possible that families were concerned about risking exposure to COVID-19 by attending a healthcare facility. [3] This anxiety could also help explain the observed increase in the proportion of patients requiring hospitalization as families may have been reluctant to seek care due to fear of contracting the virus, potentially leading to delays in consultations and more severe presentations. [20-23] A third possibility is an increase in telemedicine services, that were both more widely offered by medical professionals and more frequently used by families. [24] The significant decrease in the proportion of patients LWBS may be attributable to the overall higher acuity of the presentations to the ED, reflecting patients for whom leaving without medical assessment would be less likely, as well as lower wait times related to decreased overall volume in the ED.

Our findings need to be interpreted in light of a few limitations. First, data available for this analysis did not allow to disentangle the role and relative contribution of potential drivers of decreases in emergency visits. As such, whether there was really a delay in consultation and more severe presentations cannot be ascertained in this report. Second, our results represent visits at the two pediatric EDs in the province of Quebec and may therefore not be generalizable to other cities or countries, although the magnitude of change in ED visits is similar to other previously published studies. We were also limited to the information available in the electronic databases at each institution. For example, as the diagnostics were grouped into categories, it was not possible to tease out whether in a given category, there were increases in specific diagnoses. Finally, errors in the collection, entry and coding of data may occur, although errors in data should be evenly distributed between the two study periods.

## Conclusion

Our study demonstrated a 53.3% decrease in the median number of weekly pediatric ED visits during the first wave of the COVID-19 pandemic (March-May 2020) compared to historical control. It is important to examine and monitor trends in ED visits, including the acuity of these visits and the categories of diagnoses made, as they provide insight into changes in the population’s use of the healthcare system, acting as a first-line indicator of the presence of certain diseases in the population, as well as a barometer for access to healthcare. While decreases in pediatric ED visits may be in part attributable to reductions in communicable diseases related to social distancing and other public health measures, higher acuity visits may indicate that patients and their families had concerns about the safety of presenting to the ED and delayed or avoided seeking care. It will be important to assess whether this trend is temporary and whether there are factors underlying the reduction in the number of ED visits that should be addressed to improve access to and efficiency of pediatric emergency care.

## Data Availability

Data are available for the corresponding author on request.

## Abbreviations

CHUSJ: CHU Sainte-Justine
CTAS: Canadian Triage and Acuity Scale
ED: Emergency Department
IQR: Interquartile Range
LWBS: Left Without Being Seen
MCH: Montreal Children’s Hospital

## Acknowledgements

The authors would like to thank Ms. Tamara Perez for her help in reviewing and editing this manuscript.

## Appendices

<u>Appendix A: ICD-10 code groupings used for the categorization of MCH ED discharge diagnoses.</u>

Pediatric ED level data (ICD-10 codes)

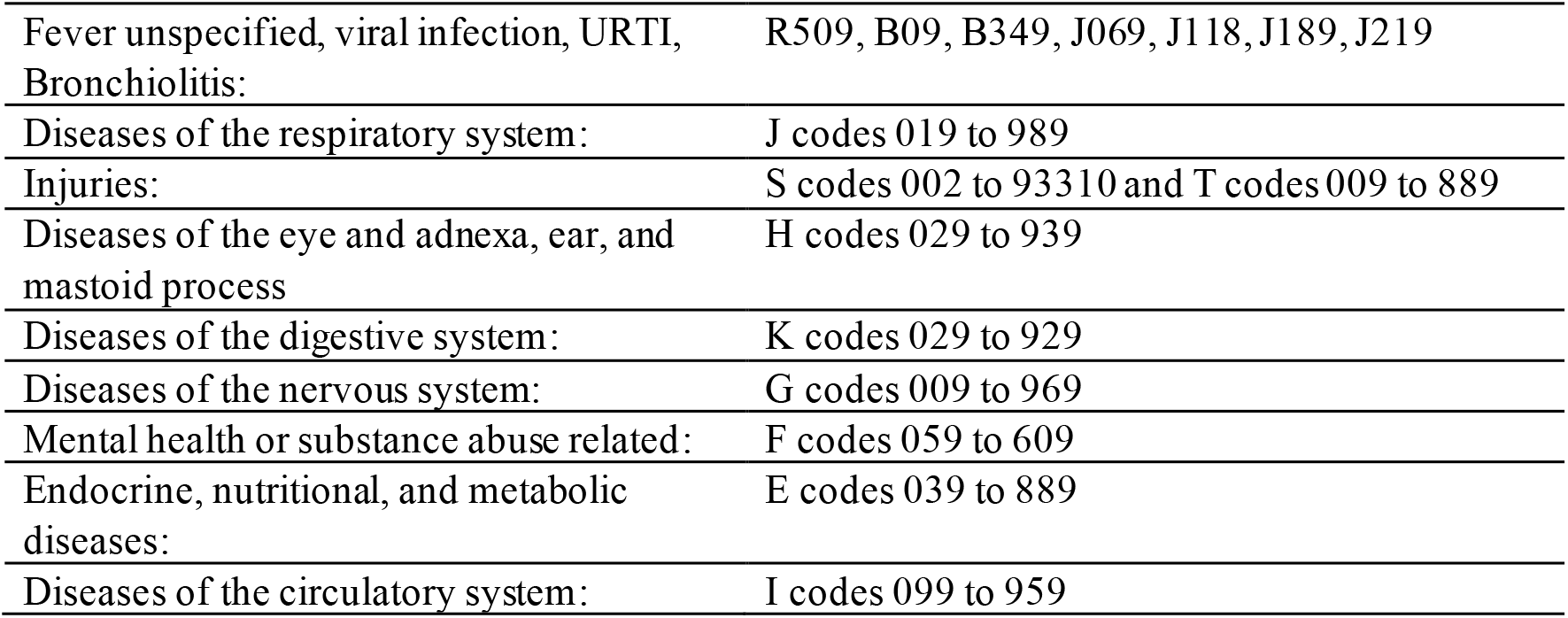

